# High burden of subclinical TB in Africa revealed from a postmortem cohort

**DOI:** 10.64898/2026.06.09.26345127

**Authors:** Gift Ahimbisibwe, Marjorie Nakibuule, Marvin Martin Ssejjoba, Marvin Joven Turyasingura, Rose Mulwana, Claire Precious Bisoboka, Kutuusa Daniel, Josephine Nabulime, Febronius Babirye, Musana Abdusalaamu Kizito, Hervé Lekuya, Akello Suzan Adakun, Daisy Nalumansi, Julius Mwesige, Aminah Nalukwago, Joseph Baruch Baluku, Robert Lukande, Irene Andia Biraro, Stephen Cose

## Abstract

Tuberculosis (TB) is increasingly recognised as a spectrum of infection and disease, yet the prevalence of viable, asymptomatic *Mycobacterium tuberculosis* (*M*.*tb*) infection remains uncertain. Subclinical Tuberculosis (scTB), defined as microbiologically confirmed *M*.*tb* infection in the absence of recognised symptoms, is under detected by symptom, sputum and imaging-based approaches. We conducted postmortem examinations of 94 adults who died from non-infectious causes, none of whom were clinically suspected of TB or reported TB related symptoms prior to death. Lung and extrapulmonary tissues were cultured for *M*.*tb*. Viable *M*.*tb* was confirmed in six individuals, corresponding to a prevalence of 6.4% (95% CI: 2.4 to 13.4%). These findings provide direct tissue-based evidence that viable, asymptomatic *M*.*tb* infection can persist beyond the reach of conventional clinical detection. Our data suggest that a biologically active reservoir of infection may exist undetected within high-burden settings, with implications for surveillance strategies aimed at TB elimination.

## Body

TB remains the leading cause of death from a single infectious agent worldwide, despite the availability of effective treatment and sustained global control efforts^1^. A major barrier to TB elimination is the inability to detect *M*.*tb* infection before the onset of clinically apparent disease. This limitation reflects an outdated binary view of TB as either latent or active. Increasingly, TB is understood to exist as a dynamic infection–disease spectrum, within which subclinical TB (scTB) represents an early stage characterised by viable infection in the absence of overt symptoms^2,3^. It is thought that individuals within this subclinical phase may contribute to transmission in high-burden settings. Reflecting this evolving understanding, recent WHO guidelines and the International Consensus for Early TB (ICE-TB) framework emphasised the importance of identifying and treating scTB before transmission risk increases and irreversible tissue damage develops^4,5^.

Despite this conceptual shift, detecting infection at this early stage remains inherently challenging, and the true prevalence of scTB remains unknown. Early infection is thought to localise in anatomically inaccessible regions of the lung that are poorly sampled by sputum-based diagnostics or bronchoscopy. Moreover, sputum is frequently unavailable and performs sub optimally in children, people living with HIV, and individuals with early infection^6-8^. Although radiological imaging is considered more sensitive, infection may precede detectable radiographic or histopathological abnormalities^9^. Therefore, current estimates that heavily rely on these approaches systematically underestimate the prevalence of early scTB^10^.

Postmortem examination offers a unique opportunity to overcome these limitations by providing direct access to lung tissue, deeper bronchoalveolar lavage compartments, and lymphoid organs that are inaccessible and cannot be comprehensively sampled in living individuals. Importantly, this facilitates the detection of viable *M*.*tb* even in the absence of macroscopic or classical pathological disease in inaccessible sites. Here, we use a postmortem approach to investigate *M*.*tb* infection in individuals admitted for acute trauma or other non-infectious emergencies at the Surgical Emergency Unit (SEU) of Mulago National Referral Hospital, and who were not clinically suspected of TB. Recruitment at the SEU is high; we have previously reported a 94% consent rate of eligible Next-of-Kin (NoK) on this ward^11^.

Between January 2021 and August 2025, 115 deceased individuals aged ≥18 years were assessed for eligibility; all had undergone routine clinical evaluation in the SEU and had participant records available. Six individuals were excluded due to known comorbidities (four with diabetes and two with asthma), leaving 109 eligible individuals without known chronic medical conditions or clinically identified chest trauma. Of these 109 subjects, the NoK of 101 individuals consented to participate. Among these, NoK interviews and clinical record review established that 98 individuals had no reported TB-related symptoms prior to death, while three had symptoms suggestive of TB and were excluded. A full-body postmortem examination was subsequently performed to establish cause of death and identify conditions missed during routine clinical assessment. Four additional individuals were excluded (three with previously undiagnosed malignancies and one with lung trauma not identified clinically), resulting in a final cohort of 94 participants.

At postmortem examination, 93 of 94 subjects had normal lung parenchyma, pleura, and pulmonary vasculature, with no macroscopic lesions suggestive of TB disease. One subject exhibited visible pulmonary lesions consistent with TB and was retained in the study as the NoK (subject’s son who was living with him) reported no signs or symptoms of TB prior to the father’s death. Histological assessment of lung tissue was otherwise unremarkable in the remaining subjects, with no microscopic features suggestive of TB. We assessed lung and selected extrapulmonary tissues for microbiological evidence of viable *M*.*tb* using MGIT liquid culture. *M*.*tb* was confirmed by positive ZN stain and MPT64 antigen detection assay, a specific marker for the *M*.*tb* complex. In cases where ZN was positive and the MPT64 antigen detection assay negative, we performed GeneXpert MTB/RIF testing to confirm MTBC. The combination of these strategies enabled the detection of early, low-burden infection in the absence of classical pathology, while also identifying individuals with established pathological TB disease without overt signs or symptoms, and that would otherwise remain unsuspected.

Viable *M*.*tb* was detected in six individuals (Table 1; Supplementary Table 1). Among these, Subjects 1–3 demonstrated a relatively short Time to MGIT culture Positivity (TTP), consistent with a high lung bacterial burden, and all three were ZN-positive prior to culture. One of these subjects had macroscopic pulmonary pathology at postmortem. In contrast, Subjects 4–6 demonstrated prolonged TTP consistent with a low-burden infection. In these individuals, ZN staining was negative prior to culture. Notably, two of these three low-burden cases had indeterminate T-SPOT.*TB*^®^ results, suggesting limited or absent detectable peripheral immune responses despite the presence of viable bacteria (Table 1).

**Table 1.** Patient demographics. Brief characteristics of the six scTB subjects identified from the SEU^a^.

|  | S1 | S2 | S3 | S4 | S5 | S6 |
| --- | --- | --- | --- | --- | --- | --- |
| Sex | M | M | M | M | M | M |
| HIV status | Negative | Negative | Negative | Negative | Positive | Positive |
| COD after PM | Head injury following blunt force trauma | Head injury secondary to blunt force trauma | Severe head injury Pulmonary TB confirmed | Severe head injury | Severe head injury | Severe head injury |
| T-SPOT.*TB*<sup>®</sup> Result | Indeterminate | Indeterminate | Positive | Positive | Positive | Positive |
| ZN<sup>b</sup> Staining result, before culture | Positive | Positive | Positive | Negative | Negative | Negative |
| ZN Tissue<sup>c</sup> | Lung | BAL<sup>d</sup> | BAL, Lung | Not Applicable<sup>e</sup> | Not Applicable | Not Applicable |
| Histology | Normal<sup>f</sup> | Normal | Granulomatous inflammation consistent with TB | Normal | Normal | Normal |
| MGIT<sup>g</sup> Result | Positive *M.tb* culture | Positive *M.tb* culture | Positive *M.tb* culture | Positive *M.tb* culture | Positive *M.tb* culture | Positive *M.tb* culture |
| MGIT Tissue<sup>h</sup> | Lung, HLN<sup>i</sup> | BAL | BAL, Lung | BAL | BAL | Lung |
| TTP<sup>j</sup> (Days) | 6, 23 | 16 | 12, 20 | 31 | 35 | 39 |
| ZN Staining result, after culture | Positive | Positive | Positive | Positive | Positive | Positive |
| ZN Tissue<sup>k</sup> | Lung, HLN | BAL | BAL, Lung | BAL | BAL | Lung |
| MPT64<sup>l</sup> | Negative | Negative | Positive | Positive | Positive | Positive |
| MPT64 Tissue<sup>m</sup> | Not Applicable | Not Applicable | BAL, Lung | BAL | BAL | Lung |
| GeneXpert<sup>n</sup> | Positive | Positive | Not Performed | Not Performed | Not Performed | Not Performed |
| GeneXpert Tissue<sup>o</sup> | Lung, HLN | BAL | Not Applicable | Not Applicable | Not Applicable | Not Applicable |

<sup>a</sup>SEU, Surgical Emergency Unit
<sup>b</sup>ZN, Ziehl-Neelsen
<sup>c</sup>ZN Tissue, Tissue(s) that were positive for ZN staining directly *ex-vivo* before culture
<sup>d</sup>BAL, Bronchoalveolar Lavage
<sup>e</sup>Not Applicable, assay was either not performed or result was negative
<sup>f</sup>Normal, Preserved tissue architecture without granulomatous inflammation, necrosis, or cavitation consistent with TB
<sup>g</sup>MGIT, Mycobacterial Growth Indicator Tube
<sup>h</sup>MGIT Tissue, Tissue(s) that were positive for MGIT
<sup>i</sup>HLN, Hilar Lymph Node
<sup>j</sup>TTP, Time to MGIT culture Positivity
<sup>k</sup>ZN Tissue, Tissue(s) that were positive for ZN staining directly after culture
<sup>l</sup>MPT64, MPT64 antigen assay result after culture
<sup>m</sup>MPT64 Tissue, Tissue(s) that were positive for MPT64 antigen assay after culture
<sup>n</sup>GeneXpert, GeneXpert MTB/RIF assay result after culture

Two of the six subjects were HIV positive (Table 1). Although viral load measurements and antiretroviral treatment histories were unavailable, flow cytometry analysis demonstrated CD4 T cell counts comparable to those of HIV-negative subjects, suggesting preserved immune profiles at the time of death (Supplementary Figure 1). Overall, this study anchors scTB to the direct detection of viable bacilli in tissue in the absence of reported signs or symptoms. The infection variability across our six subjects supports a graded infection disease continuum in which viable bacilli may precede or exist independently of visible tissue destruction.

In this study, 6 individuals met the definition of subclinical TB, defined as culture-confirmed viable *M*.*tb* infection in the absence of TB symptoms as defined by the WHO, corresponding to a prevalence of 6.4% (95% CI: 2.4–13.4%). Our estimate is lower than that reported in TB household contact studies, or studies that use symptom screening^10,12^,13. This is not surprising since such studies/individuals are known to have a higher *M*.*tb* exposure risk. Our study is not a household contact study and is more reflective of a community-based programme. The current best estimate from a community-based study in South Africa reported a scTB prevalence of 0.6%, 10 times lower than the prevalence found here^12^. By systematically sampling lung and extrapulmonary tissues and confirming viable bacilli by culture, our study identifies infection states that may not be captured within conventional sampling methods used in community screens. In addition, community screening approaches depend heavily on symptom reporting, sputum availability, and imaging abnormalities, each of which is insensitive for detecting early infection^10,14^.

By integrating clinical record review, structured verbal autopsy, predefined systematic tissue sampling, histopathological assessment, and culture-based confirmation, we demonstrate that asymptomatic *M*.*tb* infection can persist beyond what is captured by symptom-led, sputum-based, or radiology-guided screening strategies. We report a scTB prevalence of 6.4%, 10 times higher than any previous community-based screening study, reflecting a previously unknown and extremely high burden of asymptomatic TB infection in the community^12^. Our study has several limitations. The cross-sectional postmortem design precludes inference regarding progression, duration, or transmissibility of disease. The sample size was modest, and the cohort was not randomly sampled, resulting in wide confidence intervals around prevalence estimates. Trauma victims may not perfectly represent the general population with respect to TB risk factors. Despite these limitations, our findings provide the proof-of-principle that scTB can be identified in the absence of classical pathology and across a range of bacillary loads. These findings reinforce a spectrum-based understanding of TB and highlight the need for diagnostic tools capable of identifying infection before overt disease develops, thereby expanding opportunities for earlier intervention. More importantly, our data show a previously undiscovered and worryingly high burden of subclinical *M*.*tb* infection prevalence in an African community setting.

## Data Availability

All data produced in the present study are available upon reasonable request to the authors

## Supplementary Materials and Methods

### Ethics

This study obtained ethical approval from five ethics bodies: the Makerere University School of Biomedical Sciences Research & Ethics Committee (SBS-REC-721), the Mulago National Referral Hospital Ethics Committee (MHREC 1849), the Kiruddu National Referral Hospital School of Biochemical Research and Ethics Committee (CRD/ADMIN/120/1), the Uganda National Council of Science and Technology Ethics Committee (HS703ES), and the London School of Hygiene and Tropical Medicine Ethics Committee (22922).

### Consent

Post-mortem examination and tissue collection for research were conducted in accordance with Ugandan legal provisions governing post-mortem examination and the use of human tissues in research. The Inquests Act (Chapter 13) of Uganda provides statutory authority for postmortem examinations to establish cause of death, and the Uganda Health Act allows postmortem examinations with consent from the Next-of-Kin (NoK). Consequently, consent was sought from the NoK (spouse, adult child, parent, sibling, or other close adult relative) as prescribed by Ugandan law.

Trained grief counsellors sensitively approached the NoK to obtain informed consent for post-mortem examination, donation of tissues for medical research, storage of samples for future use, genetic studies, and access to previous medical records. A case record form (CRF) was completed for each participant, serving both to document reasons for consent or decline and to perform a verbal autopsy, capturing demographics, clinical history, and potential TB-related symptoms from the NoK.

### Participants

Deceased road traffic accident subjects were recruited from the Surgical Emergency Unit (SEU) at Mulago National Referral Hospital (MNRH), Kampala, Uganda, between January 2021 and August 2025.

We applied a two-step inclusion process:

#### Step 1 – Ward eligibility

Deceased individuals aged ≥18 years who had undergone a full clinical assessment in the SEU and had participant records available were considered. Participant records were reviewed alongside verbal autopsy interviews with NoK to assess comorbidities and TB-related symptoms. Individuals with known chronic medical conditions (including diabetes, liver disease, kidney disease and asthma), reported TB-related symptoms prior to death (as defined by the WHO), or clinically identified chest trauma were excluded^1^. NoK of eligible individuals were approached for consent, and only those for whom consent was obtained were included.

#### Step 2 – Post-mortem eligibility

A full-body postmortem examination was performed by the study pathologist to establish the cause of death and identify conditions not detected during routine clinical or laboratory evaluation. Individuals with previously unrecognised underlying conditions, including malignancies, or evidence of lung trauma not identified clinically, were excluded from this sub - study analysis.

### Tissues and sample collection

All lung sections were systematically examined for macroscopic TB lesions. Histopathology was performed to identify any microscopic evidence of TB. Following organ examination, samples were collected in the following order: arterial blood from the carotid artery into heparin tubes (arteries were preferred due to postmortem venous collapse), bronchoalveolar lavage (BAL) washes of both left and right lungs with 200 ml PBS, and tissue samples: right lung sections (superior, middle, inferior), left lung sections (superior, inferior), hilar lymph nodes (HLNs), peripheral lymph nodes (PLNs), and spleen. Solid tissues were placed in 50 ml tubes containing 20% FBS in RPMI medium, tightly capped, and transported at room temperature to the BSL3 laboratory at the MRC/UVRI & LSHTM Uganda Research Unit, where all sample processing was performed under BSL3 conditions.

### Histopathological assessment

Formalin-fixed lung tissue samples were embedded in paraffin, sectioned, and stained with haematoxylin and eosin (H&E) for evaluation of tissue architecture and pathological changes. Sections were examined by a study pathologist for features consistent with TB including granulomatous inflammation, necrosis, and cavitation. H&E staining was used to assess structural and inflammatory pathology; detection of acid-fast bacilli was performed separately using Ziehl–Neelsen (ZN) staining.

### *M*.*tb* isolation and detection

#### Recovery of viable *M*.*tb* using BACTEC MGIT 960

Isolation of viable *M*.*tb* was performed using the BD BACTEC™ MGIT™ 960 system. Approximately 5 mL of homogenised solid tissue, bronchoalveolar lavage (BAL, 100 ml) and blood (20 ml) was decontaminated with an equal volume of BD™ BBL™ MycoPrep™ reagent (Cat. No. 240863) for 30 minutes, with vortexing every 5 minutes. Each MGIT run included a negative and positive control sample with known bacterial load and time-to-positivity (TTP) to monitor the decontamination process and culture performance. Decontamination was stopped by addition of sterile phosphate-buffered saline (PBS), followed by centrifugation at 3,500 g for 15 minutes. The resulting pellet was inoculated into MGIT tubes supplemented with BBL™ MGIT™ PANTA™ antibiotic mixture (Cat. No. 245114) to suppress non-mycobacterial growth. Tubes were incubated in the MGIT 960 automated detection system for up to 42 days.

We found that extending MycoPrep decontamination from the manufacturer-recommended 15 minutes^2^ to 30 minutes, collecting tissues into RPMI medium supplemented with 1% penicillin– streptomycin, and initiating MGIT culture on the day of tissue retrieval reduced contamination (Supplementary Figure 1).

#### Confirmation of viable *M*.*tb*

The time-to-positivity (TTP) on the MGIT assay was used as an initial screen to rule out contamination. A TTP ≤ 48 hours of incubation was classified as fast growth consistent with Mycobacteria other than *M*.*tb* or other non-Mycobacterial species and was therefore excluded from further analysis. For samples with a TTP ≥ 48 hours, *M*.*tb* was confirmed by positive ZN stain and MPT64 antigen detection assay. The MPT64 antigen assay detects the MPT64 protein, a specific marker for *M*.*tb* complex. In cases where ZN was positive and the MPT64 antigen detection assay negative, we performed GeneXpert MTB/RIF testing to confirm MTBC. Only MGIT culture positive samples that were positive by ZN and either MTP64 or GeneXpert MTB/RIF were considered *M*.*tb* positive.

### Case definitions

#### scTB

scTB was defined *a priori* in line with WHO and ICE-TB frameworks as the presence of viable *M*.*tb* in any anatomical compartment, in the absence of documented TB-related symptoms prior to death. The presence of viable *M*.*tb* was confirmed by MGIT culture together with downstream species confirmation while symptom absence was ascertained using clinical records and a structured verbal autopsy, consistent with WHO symptom-based screening criteria.

PBMC and cell isolation from tissues

PBMC and cell isolation from tissues was performed as previously described ^3^.

### T-SPOT.*TB*^®^ assay

This assay was performed for SEU subjects using the T-SPOT^®^.*TB* kit (TB.300, Oxford Immunotec) to determine exposure to *M*.*tb* prior to death. Samples were run in duplicate. The procedure was performed as per insert, except for incubation time (48 hrs) and media (RPMI plus 10% FBS). Extensive testing showed that this time and media yielded the most reliable results from our deceased subjects, above that of the recommended protocol for live venous blood. Resultant spots were read using an ELISPOT reader (AID iSpot ELR08IFL) and were interpreted as per product insert.

### Flow cytometry

We assessed B cell, CD4 and CD8 T cell type frequencies of cells isolated from each of the tissues. The gating strategy for PBMC and tissues are shown in Supplementary Figure 2. Details of the panel and staining procedure have been previously described by our group^3^.

### Statistical Analysis

Statistical analyses and figure generation were performed using R (version 2024.09.0+375) and GraphPad Prism (version 9). Flow cytometry data were generated in FlowJo and imported into R and GraphPad Prism for analysis. Group comparisons were performed using the Kruskal–Wallis test, followed by Dunn’s test for pairwise comparisons with adjustment for multiple testing. Statistical significance was defined as an adjusted p-value < 0.05.

### Role of the funding source

The funder of the study had no role in study design, data collection, data analysis, data interpretation, or writing up results.

**Supplementary Figure 1.**
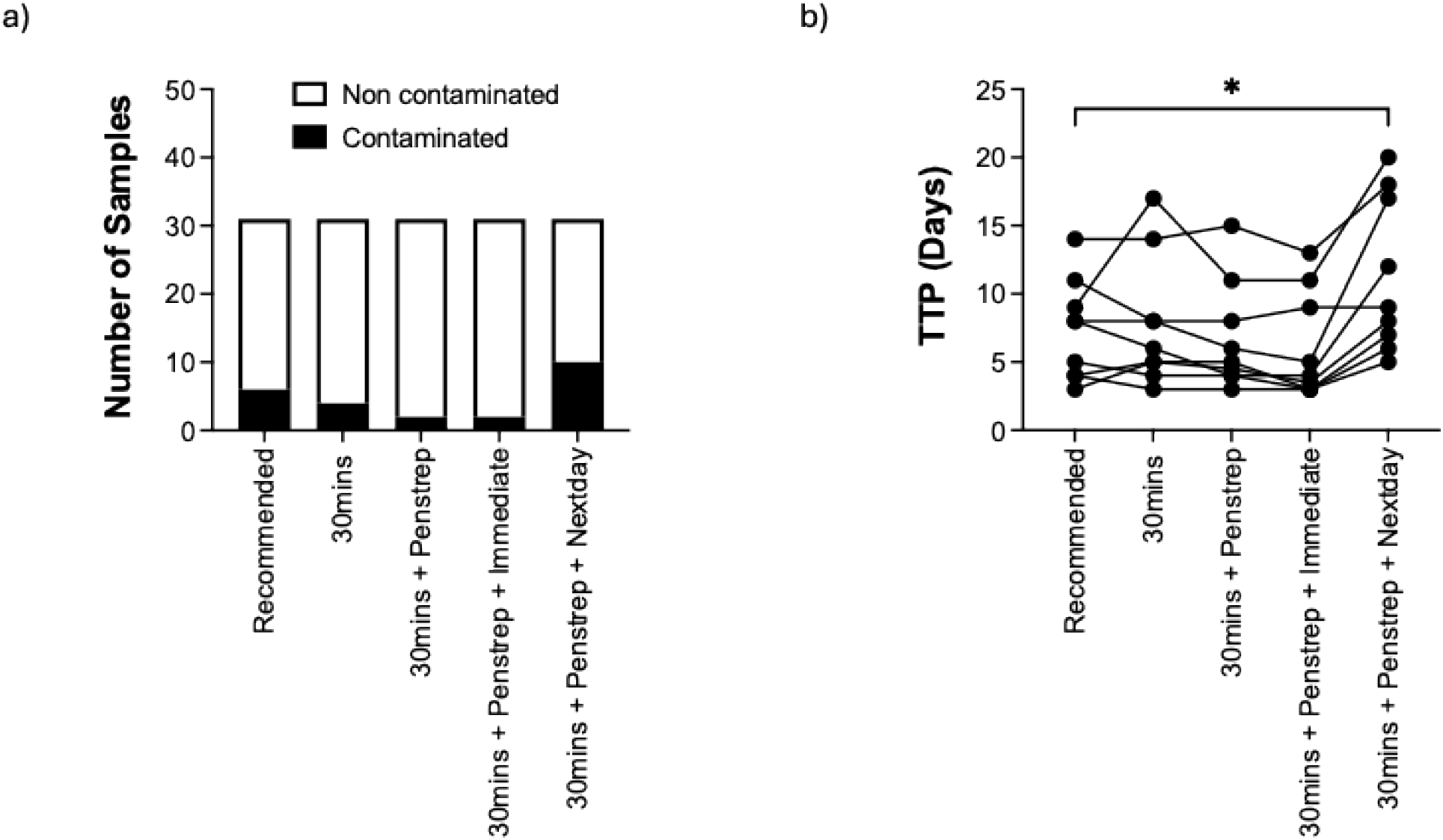
MGIT assay optimization for tissues. (a) Number of contaminated versus non-contaminated samples cultured under five conditions: (i) Recommended: manufacturer’s recommended decontamination (15 minutes), processed within 4–5 hours of arrival in the laboratory; (ii) 30mins: 30-minute decontamination, processed within 4–5 hours; (iii) 30mins + Penstrep: 30-minute decontamination with addition of penicillin–streptomycin (Penstrep) to the tissue collection media, processed within 4–5 hours; (iv) 30mins + Penstrep + Immediate: 30-minute decontamination with Penstrep, processed under 1 hour of arrival; and (v) 30mins + Penstrep + Nextday: 30-minute decontamination with Penstrep, processed the following day. (b) Time to positivity (TTP) for non-contaminated samples across the five conditions. * indicates statistical significance.

**Supplementary Figure 2.**
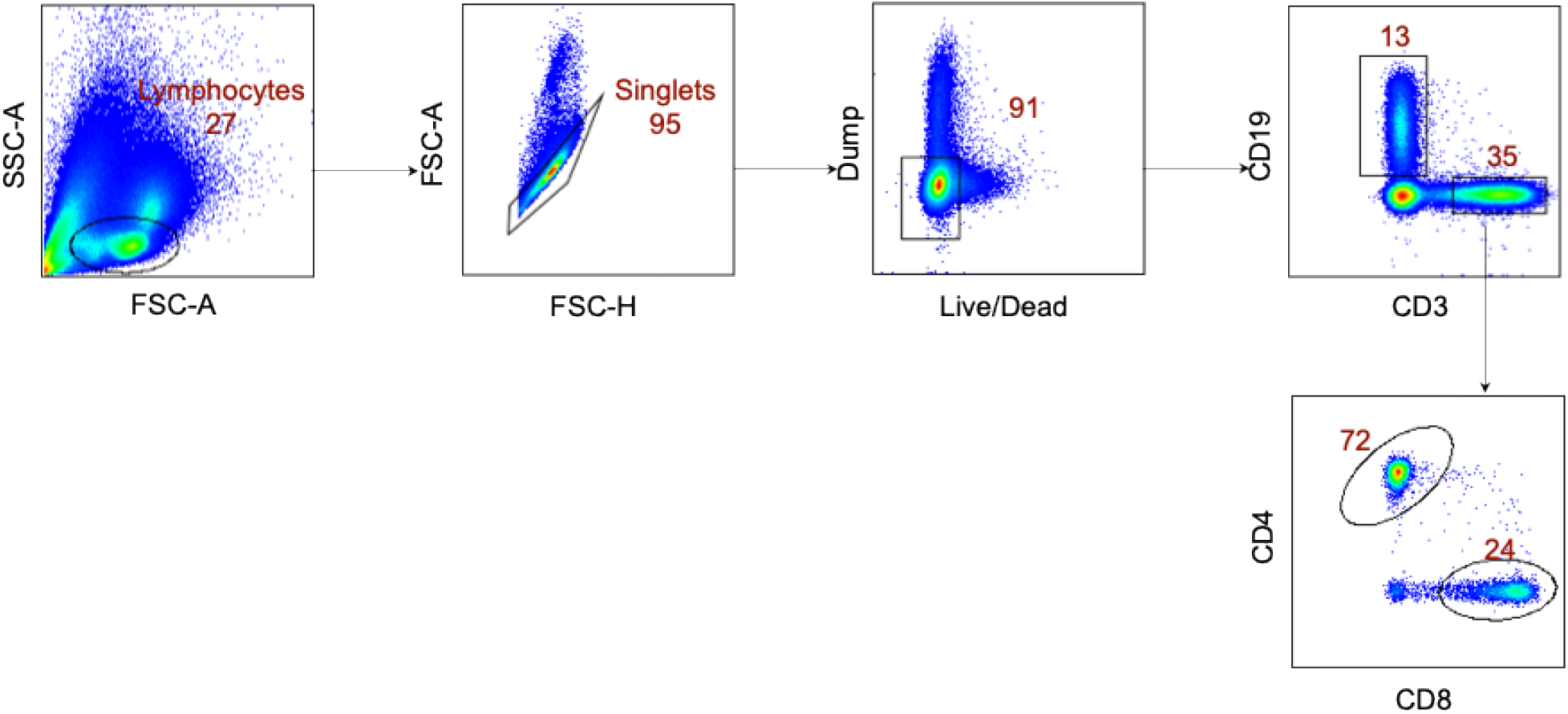
Gating strategy for flow cytometry for PBMC and other tissues. Representative plot of total cells. Cells were first gated by FSA/SSA, then singlets isolated, followed by selection of live cells and then CD3 positive cells. The dump gate, used to make the CD3 and CD19 gates cleaner, consists of CD56/CD14. CD3 positive cells were then separated based on CD4 and CD8 expression. Representative gating strategy.

**Supplementary Figure 3.**
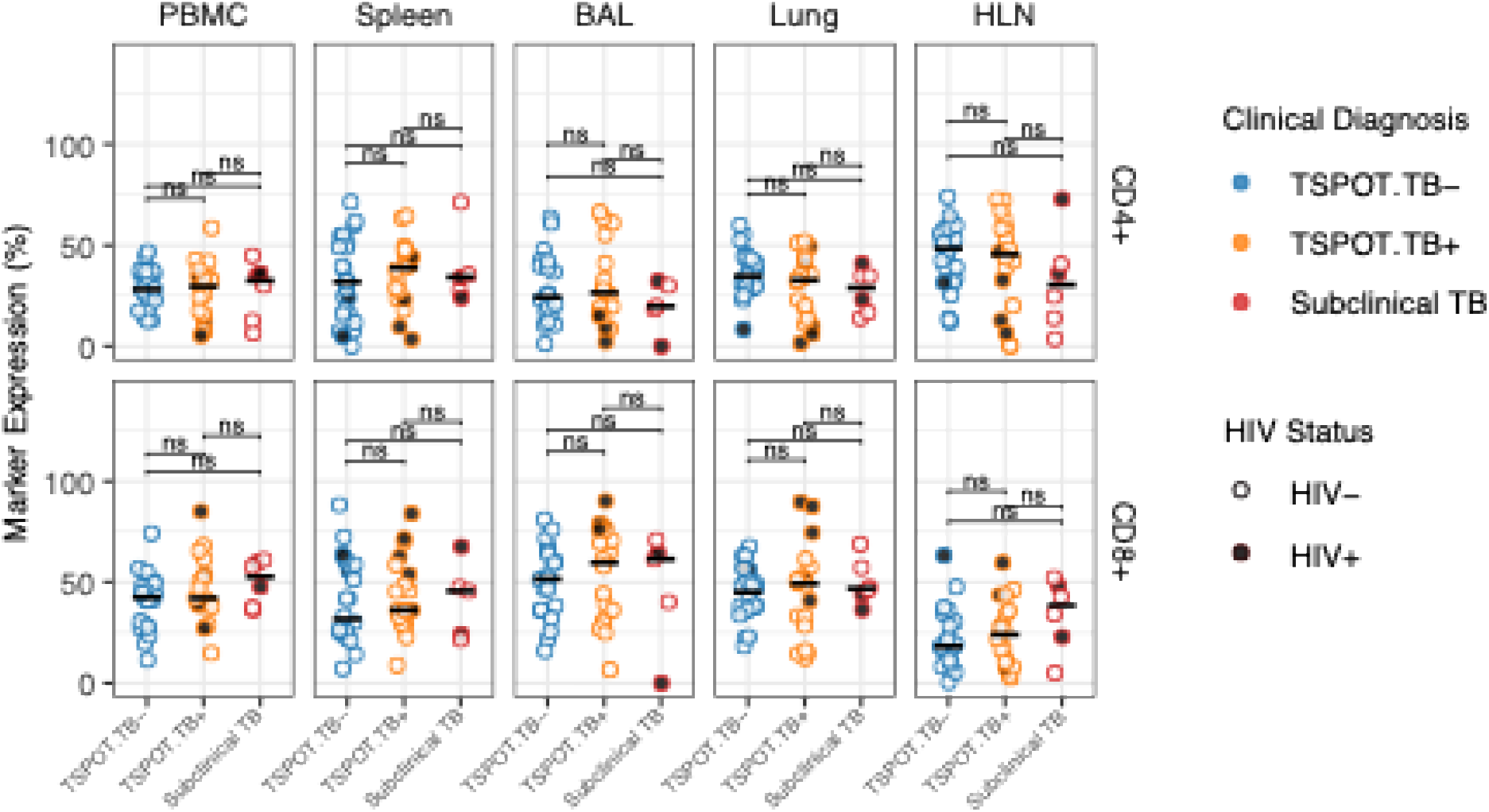
T cell frequencies across tissues in T-SPOT.*TB*^®^-defined groups and subclinical TB. Percentages of CD4^+^ and CD8^+^ T cells are shown across tissues in T-SPOT.*TB*^®^-negative (blue), T-SPOT.*TB*^®^-positive (yellow), and subclinical TB (red) individuals, stratified by HIV status. Open symbols, HIV-negative individuals; closed symbols, HIV-positive individuals.

**Supplementary Table 1.** Patient demographics. Additional characteristics of the six scTB subjects identified from the SEU.

|  | **S1** | **S2** | **S3** | **S4** | **S5** | **S6** |
| --- | --- | --- | --- | --- | --- | --- |
| Smoking/substance use | No | No | No | No | Yes | No |
| Alcohol use | No | No | No | No | Yes | Yes |
| Blood transfusion | No | No | No | No | No | No |
| HIV status | Negative | Negative | Negative | Negative | Positive | Positive |
| HIV treatment | NA | NA | NA | NA | Unknown | Unknown |
| Duration of treatment | NA | NA | NA | NA | Unknown | ~10years |
| Previous TB episode | None | None | None | None | Unknown | None |
| Type of Injury suffered | Trauma | Trauma | Trauma | Trauma | Trauma | Trauma |
| Anatomical location of injury | Head | Head | Head | Head | Head | Head |

NA<sup>a</sup> - Not Applicable

### Contributors

Gift Ahimbisibwe (AG): Formal analysis, Methodology, Project administration, Visualization, Investigation, Data curation, Writing – original draft.

Marjorie Nakibuule (MN), Marvin Martin Ssejjoba (MMS): Data curation, Methodology, Project administration, Writing – review & editing.

Rose Mulwana (RM): Methodology, Project administration, Writing – review & editing.

Claire Precious Bisoboka (CPB), Josephine Nabulime (JN), Marvin Joven Turyasingura (MJT), Febronius Babirye (FB), Musana Abdusalaamu Kizito (MAK), Daisy Nalumansi (DN), Kutuusa Daniel (KD), Julius Mwesige (MJ), Aminah Nalukwago (AM): Methodology, Writing – review & editing.

Hervé Lekuya (HL), Akello Suzan Adakun (ASA), Robert Lukande (RL), Joseph Baruch Baluku (JBB), Irene Andia Biraro (IAB): Supervision, Methodology, Writing – review & editing.

Stephen Cose (SC): Conceptualization, Funding acquisition, Supervision, Validation, Visualization, Investigation, Software, Resources, Writing – review & editing.

All authors had full access to all the data in the study and had final responsibility for the decision to submit for publication. AG, MN, MMS, and SC accessed and verified the data.

## Conflict of Interest Statements

We declare no competing interests.

## Acknowledgements

This work was funded by an NIH Contract (75N93019C00070). It was conducted at the MRC/UVRI and LSHTM Uganda Research Unit which is jointly funded by the UK Medical Research Council, part of UK Research and Innovation (UKRI) and the UK Foreign, Commonwealth and Development Office (FCDO) under the MRC/FCDO Concordat agreement and is also part of the EDCTP2 programme supported by the European Union. We are grateful to the administration of Mulago National Referral Hospital for their enthusiasm and cooperation in this research. We extend our utmost gratitude to the bereaved families who gave permission to enrol deceased relatives into the study. We also acknowledge the outstanding efforts of the clinical team who made this study possible.

## Notes

### Competing Interest Statement

The authors have declared no competing interest.

